# *MUC5AC* genetic variation is associated with tuberculosis meningitis CSF cytokine responses and mortality

**DOI:** 10.1101/2022.07.31.22278236

**Authors:** Michelle C. Sabo, Nguyen T.T. Thuong, Xuling Chang, Edwin Ardiansyah, Trinh T.B. Tram, Hoang T. Hai, Ho D.T. Nghia, Nguyen D. Bang, Sofiati Dian, A. Rizal Ganiem, Vinod Kumar, Zheng Li, Martin Hibberd, Chiea Chuen Khor, Guy E. Thwaites, Dorothee Heemskerk, Arjan van Laarhoven, Reinout van Crevel, Sarah J. Dunstan, Javeed A. Shah

## Abstract

**Rationale:** Lung mucins are an understudied component of the mucosal immune response and may influence tuberculosis pathogenesis and outcomes.

**Objectives:** To assess if variants in lung mucins *MUC5B* and *MUC5AC* are associated with *Mycobacterium tuberculosis* immune responses, susceptibility, and outcomes.

**Methods:** We characterized four haplotype tagging single nucleotide polymorphisms (SNPs) in *MUC5B* and *MUC5AC* for association with log_2_ TNF concentrations in cerebral spinal fluid (CSF) from TBM patients. SNPs associated with CSF TNF concentrations were carried forward for analyses of pulmonary and meningeal TB susceptibility and TBM mortality.

**Measurements and Main Results:** *MUC5AC* SNP rs28737416 T allele was associated with lower CSF concentrations of TNF(p=1.8*10^−8^) and IFNγ(p=2.3*10^−6^), and higher TBM, but not pulmonary TB, susceptibility (OR 1.24, 95% confidence interval 1.03, 1.49; p=0.021). Mortality from TBM was higher among participants with the rs28737416 T/T and T/C genotype (35/119, 30.4%) versus the C/C genotype (11/89, 12.4%; log-rank p=0.005) in a Vietnamese cohort (N=211). This finding was confirmed in an independent Vietnamese validation cohort (N=87; 9/87, 19.1% vs 1/20, 2.5%; log-rank p=0.02) and an Indonesian validation cohort (N=468, 127/287, 44.3% vs 65/181, 35.9%, log-rank p=0.06).

**Conclusions:** The *MUC5AC* rs28737416 T/T and T/C genotypes were associated with higher susceptibility and mortality from TBM and lower CSF concentrations of TNF and IFNγ compared to the C/C genotype, suggesting that *MUC5AC* contributes to immune changes that influence TBM outcomes.

## INTRODUCTION

Tuberculosis (TB) leads to over one million deaths annually (1). The clinical phenotype of TB disease is highly heterogenous, ranging from asymptomatic to severe disseminated infection and death (2). Tuberculosis meningitis (TBM) is the deadliest form of TB and leads to death or neurologic sequelae in over half of affected individuals (3). However, the factors that predispose individuals to symptomatic TB and TBM susceptibility and mortality are incompletely understood.

Lung mucins are complex glycoproteins on the surface of airway epithelia that inactivate potential pathogens and modulate immune signaling (4, 5). The two gel-forming mucins in the lungs, *MUC5B* and *MUC5AC*, are encoded by adjacent genes on chromosome 11p15.5 and are essential for lung health (4, 6, 7). Mice and humans with genetic mutations lacking *MUC5B* develop severe pneumonia and upper airway obstruction due to impaired mucociliary clearance, whereas overexpression leads to pulmonary fibrosis (8-11). Similarly, *MUC5AC* is strongly induced by inflammation and influences pathogen clearance, asthma pathogenesis, and may contribute to TB immune escape during helminthic co-infection by altering inflammatory responses (12-16). Therefore, mucins may influence initial TB infection by altering mucociliary clearance and initial infection of alveolar macrophages or TB dissemination by altering dynamics of inflammation. However, the role of mucins in TB pathogenesis is unknown.

Multiple lines of evidence demonstrate that host genetics influence TB risk (17). Prior genetic association studies have generally focused on immune recognition and response mediators. Variation in Toll-like receptor (TLR) 2, TLR9, mannose binding protein (MBP), leukotriene A4 hydrolase (LTA4H), and toll interacting protein (TOLLIP), have been associated with TBM susceptibility (18-22). Similarly, LTA4H and CD43 variants have been linked to higher risk of mortality from TBM (23, 24). However, the influence of genetic polymorphisms in non-immune factors produced in the lung, such as mucins, on immune function, TB susceptibility, or extrapulmonary spread are not known.

Genetic studies provide a powerful tool for examining how novel genes, such as mucin genes, influence TB pathogenesis. Evaluation of immune phenotypes that correlate with disease outcomes may detect novel variants associated with TB susceptibility and mortality. Studies of genetically regulated cytokine responses in Bacillus Calmette-Guerin (BCG)-specific T cells, dendritic cells, and peripheral blood mononuclear cells (PBMCs) have identified potential TB susceptibility factors (21, 25-27). Variants that influence functional responses in tissues of interest represent attractive secondary traits that can be correlated with TB susceptibility, and these correlations may provide insight into genetic mechanisms of disease susceptibility (25)(28). Because CSF TNF concentrations have been linked with TBM outcomes and pathogenesis (23, 29), we evaluated variants in the mucin gene region for association with TNF and other CSF cytokines, followed by evaluation of candidate susceptibility SNPs with tuberculosis outcomes, including pulmonary TB and TBM susceptibility, morbidity, and mortality.

## METHODS

### SNP selection

Genotyping and single nucleotide polymorphism (SNP) quality control (QC) are described in the **Supplementary Methods**. SNPs within and around (± 10kb), the *MUC5B* and *MUC5AC* gene region were identified using a haplotype-tagging approach from a genome-wide SNP dataset of TBM cases (N=407) and primary angle closure glaucoma (PACG) controls (N=1,139) (21, 30). Four SNPs were selected based on linkage disequilibrium (LD, r^2^ < 0.1).

As a screening step, the four selected SNPs were evaluated for associations with log_2_ concentrations (pg/mL) of TNF in cerebral spinal fluid (CSF) collected at enrollment from 154 participants in the discovery cohort (defined below). Cytokine concentrations and CSF leukocytes were measured and compared using standard statistical methods (see **Supplementary Methods**) (31). Only SNPs associated with significant changes in log_2_ TNF CSF concentrations were carried forward in further analyses. The association between SNPs of interest and CSF cytokine concentrations was confirmed using data from validation cohort 2 (N=468, defined below) as described in the **Supplementary Methods**.

### Evaluation of TB Outcomes

The association between SNPs of interest and TBM status was assessed between TBM cases (N=407) and PACG controls (N=1,1339) (21, 30) using logistic regression based on an additive genetic model (AA versus [vs] Aa vs aa), with adjustment for the first three principal components (PCs) using plink v1.9 (32). Analyses were repeated using cases of pulmonary TB as described in the **Supplementary Methods**.

The association between *MUC5AC* SNP rs28737416 and TBM morbidity and mortality was analyzed in a discovery cohort consisting of participants enrolled in a study of intensified therapy (including dexamethasone) in Vietnam (ISRCTN61649292) for whom genotyping data were available (N=211, all HIV-1 seronegative) (33). The association between rs28737416 and time to death due to TBM (33) were evaluated using Kaplan-Meier estimates and compared by log-rank testing (survival package version 3.2-13, R version 4.0.2). Hazard ratios were calculated using the Cox proportional hazards model. Potential confounders, including i) weight; ii) duration of illness; iii) age; and iv) Glasgow coma score (GCS) were evaluated for associations with the outcomes of interest; those with a p-value ≤0.2 were used in adjusted analyses.

Survival analyses were repeated in two independent cohorts. The Vietnam validation cohort included 87 participants enrolled in a randomized trial in Vietnam to assess TBM survival with dexamethasone therapy (34). All participants with genotyping data available were HIV-1 seronegative. For consistency with the Vietnam discovery cohort, only participants who received dexamethasone were included. The Indonesia validation cohort included 418 HIV-1 seronegative and 50 HIV-1 seropositive participants enrolled in a prospective cohort study of TBM in Indonesia, all of whom received dexamethasone (35, 36).

Information on censoring and missing data are in the **Supplementary Methods**. All participants provided written, informed consent. Country and institution specific ethical approvals were obtained as described (21, 30, 33-35).

### Evaluation of expression quantitative trait loci (eQTLs)

The association between SNPs and mucin mRNA expression in lung tissue was evaluated using the Genotype-Tissue Expression Project (GTEx) eQTL Calculator (https://www.gtexportal.org/home/testyourown, accessed 07/27/2021; see **Supplementary Methods**) (37).

## RESULTS

### A MUC5AC promoter region SNP is associated with lower concentrations of CSF TNF and IFNγ

A total of 126 SNPs were available in the TBM genome-wide SNP dataset within and around (± 10kb) the *MUC5B* and *MUC5AC* gene region. Three tagging SNPs were identified in *MUC5B*, and one tagging SNP was identified in the *MUC5AC* promoter region (**Figure 1A**). None of the *MUC5B* or *MUC5AC* SNPs were in high LD with one another (**Figure 1B**). *MUC5AC* SNP rs28737416 was not in LD with either of two *TOLLIP* gene SNPs (rs5743854 and rs3750920) previously linked to TBM susceptibility (**Table E1**) (21, 26).

**Figure 1:**
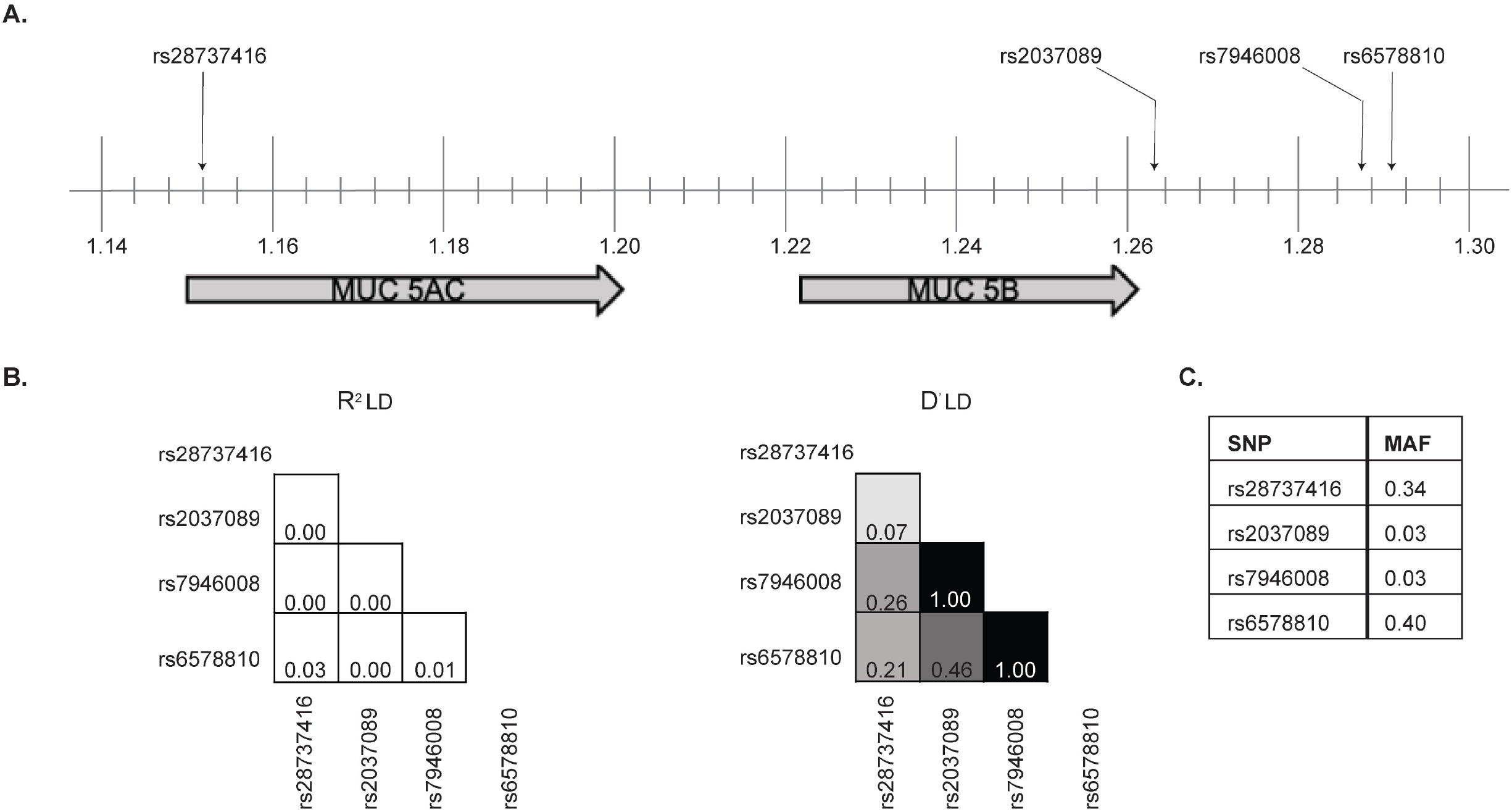
Chromosome map and linkage disequilibrium plots of haplotype-tagging SNPs identified in *MUC5B* and *MUC5AC* gene regions. **A**. *MUC5B* and *MUC5AC* genes and SNPs of interest are located on band 1 of the p arm of chromosome 11 as shown. Vertical lines represent sub-band intervals of 0.004. **B**. Linkage disequilibrium plots showing R^2^ (left) and D^’^ (right) values for tagging SNPs. The degree of shading is proportionate to the R^2^ and D^’^ values for paired SNPs. **C**. Table listing the minor allele frequency (MAF) for each SNP.

Next, we analyzed the association between haplotype tagging SNPs and TNF expression in CSF collected from participants enrolled in the Vietnam discovery cohort. Using an allelic genetic model as our primary screening analysis, the major (C) allele in *MUC5AC* SNP rs28737416 was associated with a median 2.7-fold log_2_ higher concentration of CSF TNF compared to the minor (T) allele (p=7.2 *10^−8^; **Figure 2A**). No other SNPs were associated with changes in CNS TNF cytokine concentrations. We compared alternate genetic models (additive, AA versus [vs.] Aa vs. aa genotypes and dominant, AA vs. Aa/aa genotypes) of inheritance to determine the best fitting model (**Figure 2B** and **2C**). The dominant model best fit this data, so we used this model for all future genetic analyses. Using this approach, SNP rs28737416 T/T and T/C genotypes were significantly associated with lower CSF concentrations of TNF (p=1.8*10^−8^) and IFNγ (p=2.3*10^−6^), and higher CSF concentrations of IL-2 (p=4.8*10^−6^) and IL-10 (p=0.0047) after application of a Bonferroni corrected (0.05/10 cytokines) p-value threshold of <0.005 (**Figure 2C**).

**Figure 2:**
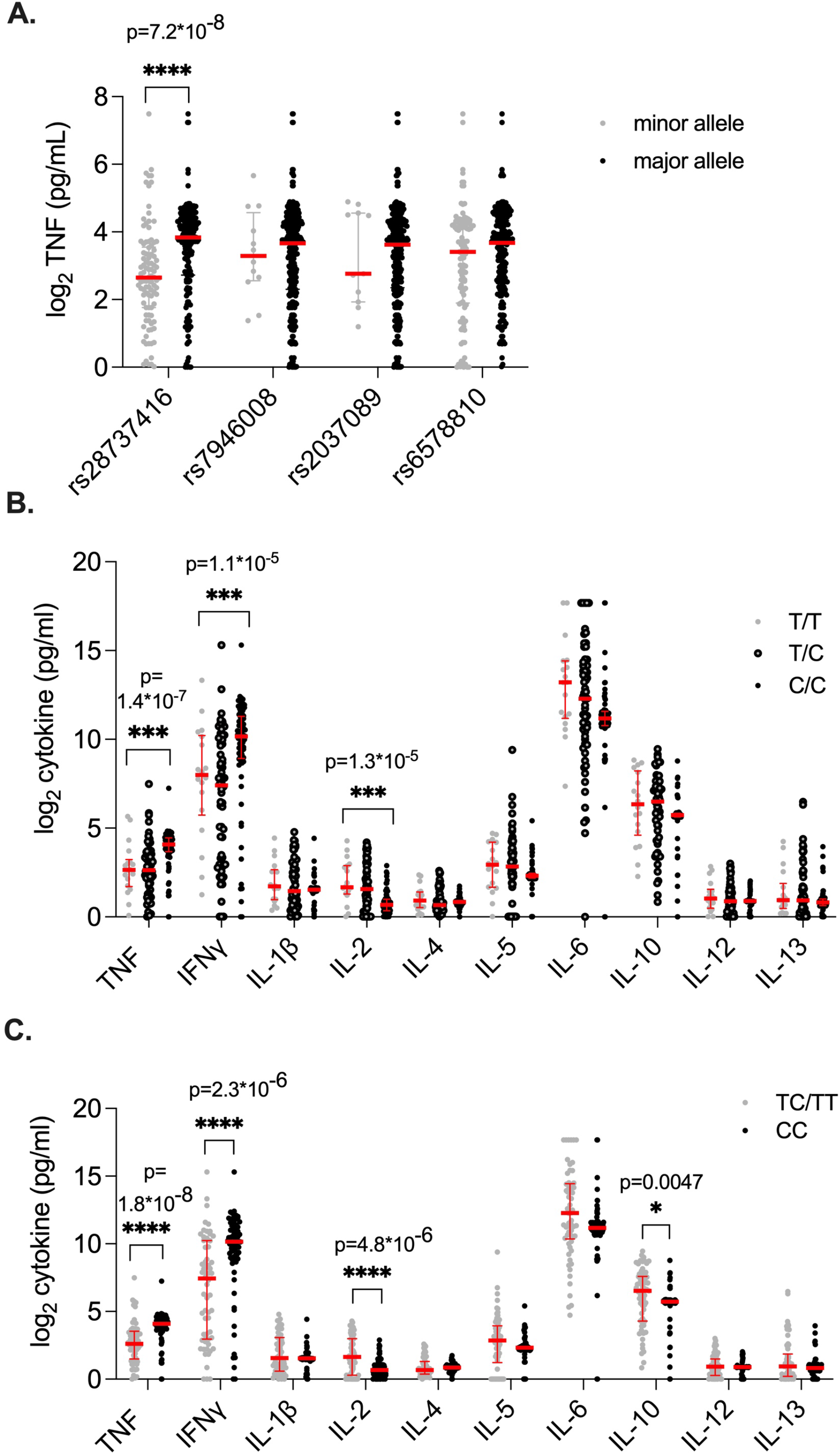
Evaluation of CSF cytokine expression in TBM patients with *MUC5B* and *MUC5AC* SNPs assessed by allelic, additive, and dominant genetic models. **(A)** Log_2_ TNF concentrations (pg/mL) measured in CSF were compared using Wilcoxon rank sum for tagging SNPs based on an allelic model (e.g., comparison of total number of alleles). For each SNP, the major allele is represented by black circles, and the minor allele is indicated by grey circles. Measured cytokine concentrations were counted twice, as all participants contribute two alleles. The association between *MUC5AC* SNP rs28737416 and additional log_2_ CSF cytokine concentrations (pg/mL) were evaluated using additive (**B**) and dominant genetic models (**C**). For the additive model, cytokine concentrations were compared between those with the T/T (grey circles), T/C (open circles), or C/C (black circles) genotypes using the Kruskal-Wallis test. In the dominant model, the composite of the T/T and T/C genotypes (grey circles) were compared to the C/C genotype (black circles) using Wilcoxon rank-sum testing. For all graphs, the median is indicated by a thick, horizontal red line, and the interquartile range is represented by thin, horizontal red lines. To adjust for multiple comparisons, a Bonferroni correction was applied to the results in B and C, and significance threshold adjusted to the level of p <0.005. P-values for cytokines with statistically significant differences in expression after Bonferroni correction are shown.

We performed PCA of cytokine concentrations to evaluate the relationship between cytokines and reduce the number of statistical comparisons. We found two uncorrelated factors, Factor 1 and Factor 2 (R^2^=0.08). Factor 1 had an eigenvalue of 5.46, explaining 74.1% of the variability in the data. Factor 2 had an eigenvalue of 1.02, explaining 13.8% variability in the data, and was influenced almost entirely by variability in TNF (loading value=0.59) and IFNγ (loading value=0.57) (see **Supplementary Results**). Factor 1 was not associated with the presence of the C/C genotype (exponentiated [exp]β=0.88, 95% CI 0.73, 1.06; p=0.19). However, Factor 2 was associated with the C/C genotype (expβ=0.63, 95% CI 0.50, 0.79; p=4.9*10^−5^), suggesting that variations in TNF and IFNγ concentrations are influenced by differences in genotype.

We validated the association of rs28737416 with TNF concentrations in the CSF collected from participants in the Indonesia validation cohort. As in the Vietnam discovery cohort, the T/T and T/C genotype of rs28737416 was associated with lower TNF concentrations (estimate [est] = -0.44, p=0.008) and IFNγ (est = -0.67, p =0.03). The relationship between expression of additional CSF cytokines and TBM in this cohort are presented in Table **E2**. These data demonstrate the generalizability of this genetic phenotype of TBM immune response across ethnically distinct populations.

Based on these preliminary observations, we evaluated the association between the candidate SNP rs28737416 and total number of CSF white blood cells (expβ=1.23, 95% CI 0.90, 1.70; p=0.19), neutrophils (expβ=1.05, 95% CI 0.83, 1.34; p=0.68), and lymphocytes (expβ=1.25, 95% CI 0.90,1.74; p=0.18) in the Vietnam discovery cohort using a dominant genotypic model. The expβ representing the mean change in cell concentration between participants with a composite of the T/C and T/T genotypes compared to participants with the C/C genotype. These data suggest that *MUC5AC* SNP rs28737416 T allele is associated with lower CSF concentrations of TNF and IFNγ in individuals with TBM, independent from total CSF cell count.

### MUC5AC SNP rs28737416 is associated with TBM susceptibility and mortality

Because rs28737416 T allele was associated with lower TNF in individuals with TBM, we further analyzed the association between *MUC5AC* SNP rs28737416 and TBM susceptibility by case-control analysis (see methods). SNP rs28737416 was associated with susceptibility to TBM (OR=1.24, 95% CI 1.03, 1.49; p=0.02) (**Table 1**), but not pulmonary TB (OR=1.11, 95% CI 0.98, 1.25; p=0.10).

**Table 1:** Association of *MUC5AC* SNP rs28737416 with diagnosis of TB meningitis and pulmonary TB.

| TB Meningitis |  |  |  |  |
| --- | --- | --- | --- | --- |
| Genotype | Controls<sup>†</sup><br>(N = 1,139) | Cases<br>(N = 407) | OR (95% CI) | P-value |
| C/C | 582 (51.3%) | 182 (45.4%) | 1.24 (1.03, 1.49) | 0.021 |
| T/C | 464 (40.9%) | 169 (42.1%) |  |  |
| T/T | 89 (7.8%) | 50 (12.5%) |  |  |
| Pulmonary TB* |  |  |  |  |
|  | Controls<sup>†</sup><br>(N = 1,270) | Cases<br>(N = 1,380) |  |  |
| C/C | 655 (51.6%) | 662 (48.0%) | 1.11 (0.98, 1.25) | 0.100 |
| T/C | 510 (40.2%) | 588 (42.6%) |  |  |
| T/T | 100 (7.9%) | 129 (9.3%) |  |  |

The association between diagnosis of TB meningitis or pulmonary TB (analyzed separately) and *MUC5AC* SNP rs28737416 was compared between cases and controls as described in the methods. The total number of individuals with major homozygous (C/C), heterozygous (T/C) and minor homozygous (T/T) genotypes are shown.
\* Genotype data were missing for 5 cases and 1 control.
<sup>†</sup> Controls were generated from the same cohort of participants with primary angle closure glaucoma.
Abbreviations: TB, tuberculosis; SNPs, single nucleotide polymorphisms; OR, odds ratio; CI, confidence interval.

Selected genes may influence progression of disease above susceptibility (24), so we next considered whether *MUC5AC* SNP rs28737416 was associated with TBM mortality in the Vietnam discovery cohort and two independent validation cohorts. Baseline characteristics for each cohort are shown in **Table 2**.

**Table 2:**
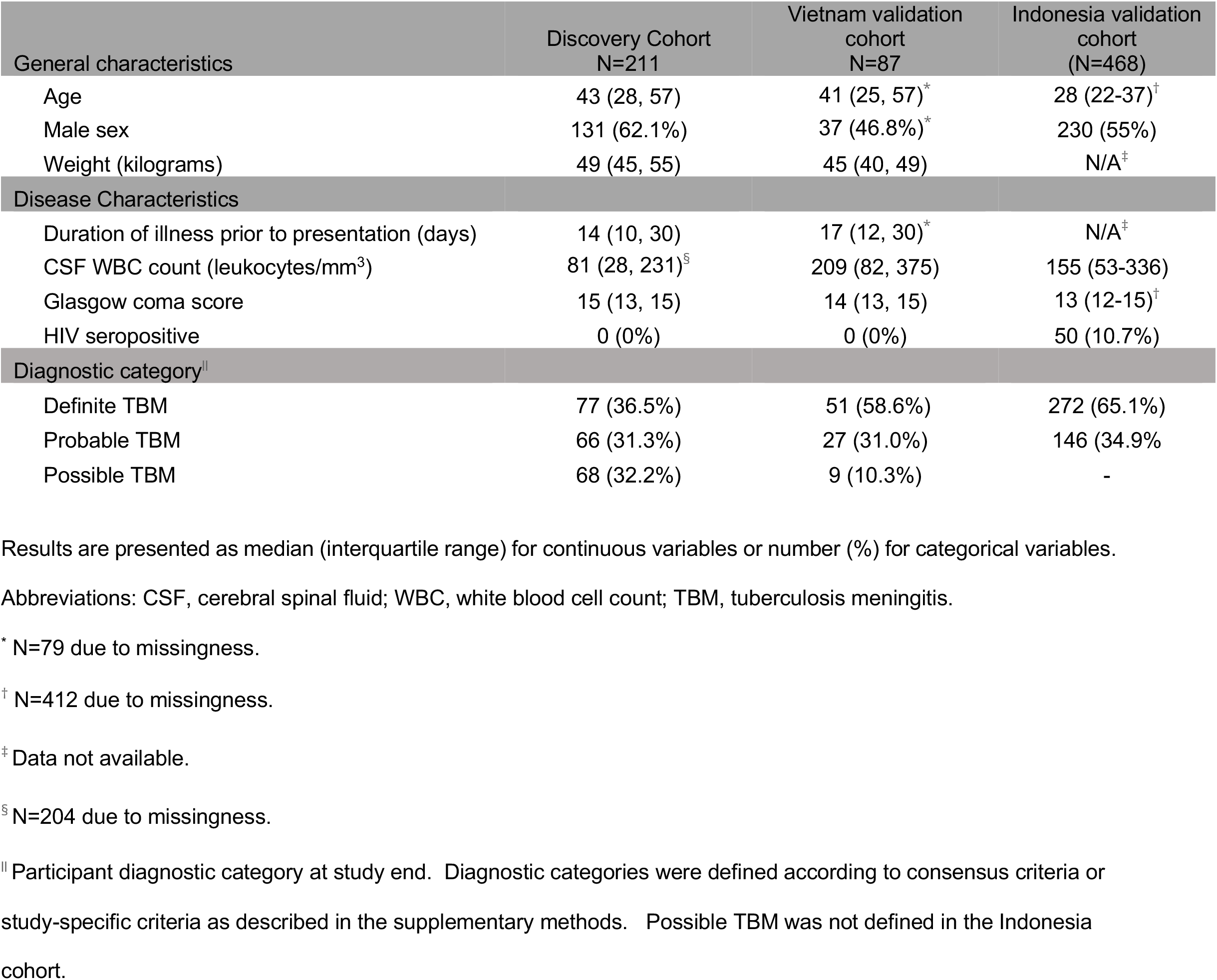
Baseline characteristics.

Survival analyses were first performed in the Vietnam discovery cohort with SNP rs28737416. We found a greater proportion of participants with T/T and T/C genotypes died (35/119, 30.4%) compared to those with the C/C genotype (11/89, 12.4%) (p=0.005) (**Figure 3A**), which corresponded to a 61% lower risk of death among participants with the C/C genotype (hazard ratio [HR] 0.39, 95% CI 0.20, 0.77; p=0.006). This association persisted after adjustment for age and GCS (adjusted HR [aHR] 0.31, 95% CI 0.16, 0.63; p=0.001). Adjustment for principal components representing genetic substructure did not alter the results (data not shown). SNP rs2873716 T allele was associated with higher risk mortality in an additive model as well (**Figure E1**).

**Figure 3:**
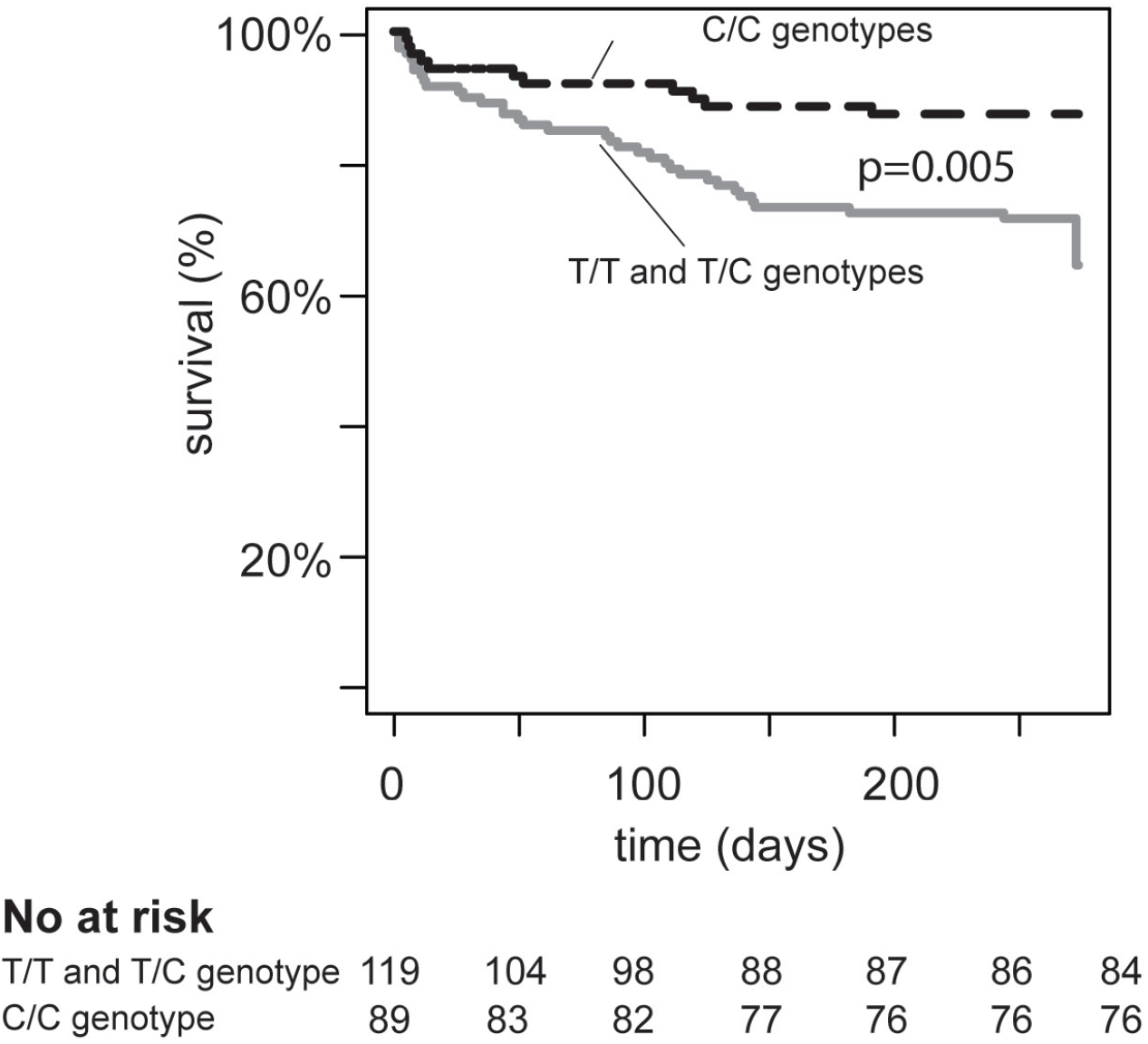
Kaplan Meier curves comparing survival in a dominant allelic model of *MUC5AC* SNP rs28737416 in the discovery cohort. Kaplan-Meier curves displaying time to death using a dominant allelic model comparing participants with the T/T and T/C genotypes to those with the C/C genotype are shown. Displayed p-values represent comparison of the survival distributions by the log-rank test.

Results were confirmed in a Vietnam validation cohort and a separate Indonesia validation cohort. In the Vietnam validation cohort, more T/T and T/C participants (9/47, 19.1%) died compared to those with the C/C genotype (1/40, 2.5%; p=0.02) (**Figure 4A**), representing an 87.1% lower risk of death from TBM (HR 0.13, 95% CI 0.02, 1.02, p=0.05). The association was similar after adjustment for age, weight, and GCS (aHR 0.13, 95% CI 0.02, 1.13; p=0.06).

**Figure 4:**
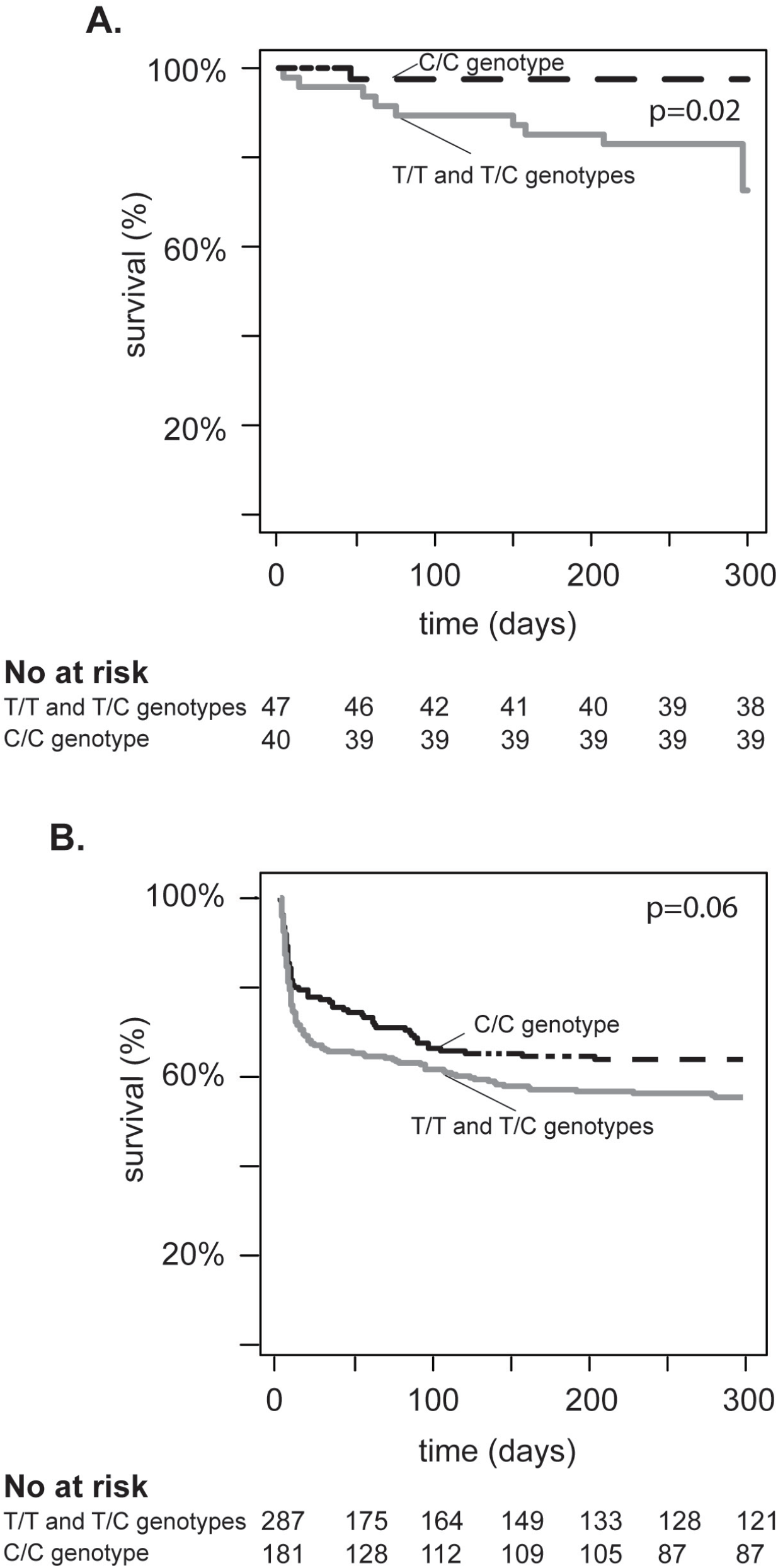
Kaplan Meier curves comparing survival in a dominant allelic model of *MUC5AC* SNP rs28737416 in the validation cohorts. Kaplan-Meier curves displaying time to death using a dominant allelic model comparing participants with T/T and T/C genotypes to those with the C/C genotype in the Vietnam validation cohort (**A**) and the Indonesia validation cohort (**B**) are shown. Displayed p-values represent comparison of the survival distributions by the log-rank test.

We measured the association between rs28737416 and TBM mortality in the Indonesia validation cohort, which included both HIV-1 seronegative (N=418) and seropositive (N=50) participants. A higher number of participants with the rs28737416 T/T and T/C genotypes died (127/287, 44.3%) compared to those with the C/C genotype (65/181, 35.9%, p=0.06) (**Figure 4B**), corresponding to a 20.1% lower risk of death among participants with the C/C genotype (HR 0.80, 95% CI 0.66, 0.99; p=0.04). However, this association was attenuated after adjustment for age and GCS (aHR=0.87, 95% CI 0.69, 1.09 p=0.22). In sensitivity analyses using only HIV seronegative participants, more participants with the T/T and T/C genotypes (106/252, 42.1%) died compared to those with the C/C genotype (56/166, 33.7%, p=0.08). The resultant risk of death was 24.8% lower among participants with the C/C genotype, and of borderline statistical significance (HR=0.75, 95% CI 0.54, 1.04; p=0.08) (**Figure E2**). Data on the proportional hazards assumptions for both the discovery and validation cohorts is included in the **Supplementary Results**. Taken together, these data suggest that *MUC5AC* SNP rs28737416 is associated with TBM susceptibility and mortality.

### SNP rs28737416 is associated with mRNA expression of MUC5AC but not MUC5B

To explore potential mechanisms for the association between mucins and TBM mortality, the association between rs28737416 and *MUC5B* / *MUCAC* mRNA expression in healthy lung tissues was assessed using public datasets (37). Participants with the T/T (0.11) genotype had significantly higher median normalized *MUC5AC* mRNA expression compared to those with the T/C (0.06) or C/C (−0.05) genotype (normalized effect size [NES] -0.14, p=0.009). There was no association between rs28737416 and *MUC5B* expression in lung tissues (NES= -0.021, p=0.67). Thus, the rs28737416 T allele is associated with higher basal *MUC5AC* expression in lung tissue, lower CSF expression of TNF and IFNγ, and higher risk of TBM mortality.

## DISCUSSION

This is the first study to link polymorphisms in a mucin gene with tuberculosis disease. Specifically, SNP rs28737416 T allele was associated with higher expression of *MUC5AC* in lung tissues, lower CSF concentrations of TNF and IFNγ in participants with TBM, and higher susceptibility and risk of death from TBM. These findings suggest that the mucosal barrier plays an important role in *Mycobacterium tuberculosis* pathogenesis.

Common variants in *MUC5AC* are associated with lung diseases, including COVID-19, and asthma (38-40). Each of these variants are in strong LD with our variant of interest in the Vietnamese population, suggesting that functional variation in *MUC5AC* may influence disease outcomes. In this study, genetic variation associated with diminished *MUC5AC* expression in the lungs of healthy individuals was protective from development of TBM. *MUC5AC* is induced during inflammation and protects against viral infection (4, 14, 15). However, over-expression of *MUC5AC* is associated with severe asthma exacerbation and ventilator-associated lung injury (13, 41). Thus, our findings fit a model where MUC5AC contributes to the pathogenesis of respiratory inflammation and infection.

How *MUC5AC* contributes to TBM mortality is unclear. It is unlikely that *MUC5AC* directly influences meningeal inflammation, as *MUC5AC* is not expressed in the central nervous system or within *M. tuberculosis*-infected myeloid cells (42). One possible explanation for the findings presented here is that over-expression of *MUC5AC* facilitates breakdown of the mucosal barrier, alveolar passage through the respiratory epithelium, and dissemination of *M. tuberculosis*. A similar mechanism of action has been proposed for the complement protein mannose binding protein (MBP), which may promote dissemination to the brain at higher concentrations (22, 43). Furthermore, *MUC5AC* may influence systemic immune responses, leading to altered *M. tuberculosis* pathogenesis. For example, *MUC5AC* variants are associated with total peripheral eosinophil count, suggesting that *MUC5AC* may play a role in conditioning systemic immune responses (38, 44). In this study, TNF and IFNγ concentrations were lower in the CSF of participants with the susceptible phenotype, while IL-2 concentrations were higher. These findings are consistent with other studies demonstrating that lower concentrations of CSF TNF and IFNγ are associated with TBM mortality among persons who receive dexamethasone therapy (23).

Based on the pattern of cytokines influenced rs28737416, *MUC5AC* may be important for the development of effective T cell responses during *M. tuberculosis* infection. Protective T cell responses against *M. tuberculosis* are generated when *M. tuberculosis* antigens are trafficked by dendritic cells from the lung to draining lymph nodes (LN), leading to T cell activation and differentiation (45, 46). Polymorphisms in *MUC5AC* may delay DC migration to the LN, which impairs immunity to *M. tuberculosis* in mice (46). Further study using methods to measure macrophage and T cell responses in the presence of *MUC5AC* polymorphisms will be required to better understand how *MUC5AC* influences TBM pathogenesis.

This study should be interpreted in the context of several limitations. First, it is possible that the susceptibility findings from the case-control analysis were detected due to another SNP (eQTL) being responsible for the change in *MUC5AC* expression. However, this appears unlikely, as rs28737416 was only associated with *MUC5AC*. Second, there is a potential for confounding due to population structure in the highly homogenous Vietnamese population. To address this potential limitation, adjustment for principal components representative of genetic substructure was performed and did not significantly alter the results. Additionally, the findings were confirmed in a separate cohort of participants in Indonesia, further suggesting genetic substructure was not influencing these results. The association in the Indonesia validation cohort was not as strong as in the Vietnamese cohorts, although the similarity in the trend in an ethnically distinct cohort provides additional evidence for an association between *MUC5AC* and TBM. The reason for this discrepancy may be due to differences in i) illness severity or time to clinical presentation across cohorts; ii) sample-sizes between cohorts; iii), linkage disequilibrium patterns across ethnicity; or, iv) environmental influence. Nonetheless, these data provide some of the strongest genetic association between a polymorphism and TBM currently. Future studies in additional cohorts and ongoing fine mapping studies may provide further insights into this modest discrepancy (23, 47, 48). Third, some participants may have been misclassified at enrollment with TBM. However, classification of TBM was performed using pre-specified criteria, which generally results in random misclassification (e.g., non-differential misclassification) and would bias away from finding an association. Fourth, although we found an association between rs28737416 and lung expression, *MUC5AC* is also expressed in the gastrointestinal tract. It is possible that TBM susceptibility is modified by changes in gut microbiota that are influenced by *MUC5AC* (47). Thus, these findings provide intriguing data for future mechanistic study.

In summary, polymorphisms in *MUC5AC* are associated with higher lung *MUC5AC* mRNA expression, lower TNF and IFNγ in the CSF, and higher risk of mortality due to TBM. These data provide evidence that respiratory mucins play an important role in TBM pathogenesis.

## Supporting information

Supplementary Results

## Data Availability

All data produced in the present work are contained in the manuscript

## ACKNOWLEDGEMENTS

We would like to thank all the participants in the parent studies used in these analyses. We would also like to acknowledge the clinical, laboratory and administrative staff at each study site for their dedication and assistance with this project. Finally, we would like to acknowledge the GTEx Project, which supports the platform on which eQTL analyses were performed. At the time of publication, the GTEx Project was supported by the Common Fund of the Office of the Director of the National Institutes of Health, and by NCI, NHGRI, NHLBI, NIDA, NIMH, and NINDS. The data used for the analyses described in this manuscript were obtained from the GTEx Portal eQTL calculator on 07/28/2021.

