## Supplementary Methods for "*MUC5AC* genetic variation is associated with tuberculosis meningitis CSF cytokine responses and mortality"

***Study subjects and tuberculosis genome wide-SNP dataset***

The Vietnam tuberculosis (TB) genome-wide SNP dataset was generated from DNA collected from participants recruited to a number of clinical studies in Ho Chi Minh City, Vietnam (1-3). Briefly, subjects with tuberculous meningitis (TBM) were recruited from two centers in Ho Chi Minh City, Vietnam: Pham Ngoc Thach (PNT) Hospital for Tuberculosis, and the Hospital for Tropical Diseases (HTD) (2, 3). Subjects with pulmonary TB were recruited from PNT hospital, as well as a network of district TB control units within Ho Chi Minh City, Vietnam, that provide directly observed therapy for treatment of TB (1). Vietnamese population control data was sourced from a GWAS of healthy adults with primary angle closure glaucoma, which has been previously described (4). The study participants were unrelated, and greater than 95% were of the Vietnamese Kinh ethnicity. Written informed consent was obtained from participants or their relatives if the participant could not provide consent (e.g., was unconscious). Individuals in the TBM group were recruited for genetics studies from 2001 to 2014 and based on the following inclusion criteria: i) age ≥ 15 years; ii) admission to the aforementioned medical centers with clinical meningitis (defined as nuchal rigidity and abnormal cerebrospinal fluid parameters); iii) a negative HIV test result; and iv) a positive Ziehl–Neelsen stain for acid-fast bacilli or *Mycobacterium tuberculosis* culture, or both, from cerebrospinal fluid (“definite TBM”). In addition to definite TBM, the cohort included subjects with “probable TBM,” defined as clinical meningitis plus at least one of the following: i) chest radiograph consistent with active TB; ii) acid-fast bacilli found in any specimen other than cerebrospinal fluid; and iii) clinical evidence of extrapulmonary TB. The pulmonary TB cohort included outpatients ≥18 years of age with no previous history of TB treatment, a negative HIV test, no evidence of miliary or extrapulmonary TB, and sputum smear with for acid-fast bacilli or culture of *M. tuberculosis* from sputum (1).

*Genotyping and Imputation:* Genotyping was performed using the Illumina HumanOmniExpress array v1.0. Standard quality control (QC) measures were applied. In brief, samples with low call-rates <95.0% (N = 23) and extremes in heterozygosity (> 3 standard deviations [SD] or < -3 SD, N = 32) were excluded. Identity-by-state measures were performed by pair-wise comparison of samples to detect sample relatedness and one sample from each pair was removed from further analysis (N = 55). Principal component analysis (PCA) was performed using the case-control dataset and the 1000 Genomes Projects (1kgP) reference populations to identify possible outliers from reported ethnicity (N=181 samples excluded).

For SNP QC, non-autosomal SNPs, SNPs with minor allele frequency (MAF) < 1.0% and low call-rates (<95.0%), and gross Hardy–Weinberg equilibrium (HWE) outliers (P < 1 × 10^−6^) were excluded. Additional autosomal SNPs were imputed with IMPUTE v2 (5) using the cosmopolitan 1000 Genomes haplotypes as a reference panel (Phase 3). SNPs with imputed information scores < 0.8, MAF < 1.0%, HWE P < 1 × 10^−7^, and non-biallelic SNPs were excluded from subsequent analyses. After QC procedures, 5,325,493 genotyped and imputed SNPs were available in the TBM GWAS dataset. Selection of SNPs of interest was then performed as described in the main methods under “***SNP Selection***.”

***Vietnam Discovery Cohort***

The Vietnam discovery cohort consisted of a subset of 211 HIV-1 seronegative participants enrolled in the 05TB study, which was a randomized controlled trial to assess the efficacy of an early, intensified treatment regimen for TBM (ISRCTN61649292) (3). Although this trial also enrolled HIV-1 seropositive participants, genotyping data were only available for participants that were HIV-1 seronegative. All participants received early, adjunctive dexamethasone therapy. Participants were enrolled based on clinical presentation, and TBM disease status was classified at study end as “definite,” “probable,” or “possible,” according to published, consensus criteria (2, 6) . Participants with unlikely TBM or an alternative diagnosis were excluded from the current analyses. Genotyping was performed using the Illumina human omniexpress_array v1.0 as described above. Censoring occurred at 275 days after randomization or at the time of the last recorded outcome. Data from 3/211 participants were not analyzed due to missingness.

***Vietnam Validation Cohort***

The Vietnam validation cohort was comprised of 87 HIV-1 seronegative participants enrolled in a randomized controlled trial to assess the effect of early, adjunctive dexamethasone on survival after diagnosis of TBM in Vietnam (2). Although this trial enrolled participants who were HIV-1 seropositive, genotyping data were only available for participants that were HIV-1 seronegative. As all participants in the discovery cohort received dexamethasone therapy, only participants in the intervention arm of the validation cohort (e.g., those that received dexamethasone) were included in these analyses. Participants were enrolled based on clinical presentation, and TBM disease status was classified at study end as “definite,” “probable,” or “possible,” as defined in the study protocol, which was conducted prior to the establishment of consensus criteria (2, 6). Participants with unlikely TBM or an alternative diagnosis were excluded from the current analyses. Genotyping was performed using the Illumina human omniexpress_array v1.0 as described above. Censoring occurred at 300 days after randomization or at the time of the last recorded outcome. Data on sex, age, and duration of illness were missing for 8/87 participants; these participants were excluded from adjusted analyses.

***Indonesia Validation Cohort***

The Indonesia validation cohort was comprised of 418 HIV-1 seronegative and 50 HIV-1 seropositive participants enrolled in a prospective cohort study of TBM at a referral hospital in Bandung, Indonesia (7). Participants were classified as having definite TBM (positive TB culture or PCR, or acid-fast bacilli on cerebral spinal fluid [CSF] gram stain) or probable TBM (CSF to blood glucose ratio of <0.5 and ≥ 5 cells/μL in participants enrolled based on clinical criteria consistent with diagnosis of TBM) at study end (7). All enrollees received treatment, including dexamethasone therapy, per current guidelines. Genotyping was performed using previously published methods (8). These methods differ slightly from those used for genotyping and imputation in the Vietnamese cohorts. In brief, samples were genotyped using a SNP chip (HumanOmniExpressExome-8 v1.0; Ilumina; San Diego, CA, USA). Samples with call rates <0.99 using standard settings on the Opticall 0.7.0 platform were excluded (9). SNPs with HWE with p<0.0001 and MAF <0.001 were also excluded. Imputation was performed using the Michigan imputation server with an Asian population reference panel (10, 11). Censoring occurred at 300 days after randomization or at the time of the last recorded outcome. Data were available to adjust for age and GCS for 412/468 participants.

***Evaluation of CSF cytokines***

Cerebral spinal fluid (CSF) was collected at enrollment in the discovery cohort (N=154) and validation cohort 2 (N=468). Cytokine protein concentrations (interleukin [IL]-1β, IL-2, IL-4, IL-5, IL-6, IL-10, IL-12, IL-13, TNF, and interferon gamma [IFNγ]) were measured from cerebral spinal fluid (CSF) from participants in the discovery cohort on a Luminex platform (Luminex, Austin TX, USA) using the R&D multiplex human cytokines kits (R&D Systems, Minneapolis MN, USA) as described (12). Cytokine concentrations in CSF collected from participants in the Indonesia validation cohort and were measured using the Olink proteomics system (Olink, Uppsala Sweden) as previously published(13).

For the Vietnam discovery cohort, log_2_ cytokine concentrations (pg/mL) were compared using the Wilcoxon rank-sum test (dominant and allelic genotypic models) and the Kruskal-Wallis test (additive genotypic model). Adjustment for multiple comparisons was performed using a Bonferroni correction (p-value threshold adjusted to <0.005). Additionally, principal component analysis (PCA) was performed and the associations between SNPs of interest and factors identified by PCA were determined using generalized linear regression with a Poisson distribution and log link with robust standard errors. To confirm that the associations seen between cytokines and SNPs of interest were not due to higher concentrations of leukocytes among individuals with TBM, the associations between CSF white blood cells and SNPs of interest was also analyzed using generalized linear regression with a Poisson distribution and log link with robust standard errors. The association between SNPs and cytokine concentrations in the Indonesia validation cohort were analyzed using linear regression with ordinary least squares estimation and a Benjamini-Hochberg correction to adjust for multiple comparisons. Analyses were performed in Stata Version 15.1 (College Station, TX, USA) and GraphPad Prism Version 9.2.0 (San Diego, CA, USA).

***Evaluation of expression quantitative trait loci (eQTLs)***

The primary output of eQTL analysis using the Genotype-Tissue Expression Project (GTEx) eQTL Calculator is the “normalized effect size” (NES). The NES can be interpreted similarly to a β coefficient generated by linear regression and represents the change in mRNA expression in persons with the homozygous minor allele genotype compared to those with the major allele genotype as listed in the GTex database. Of note, the dominant allele in the Vietnam cohorts used here was inverse to the dominant allele listed in the GTex database; thus the sign (positive or negative) reported for the NES does not accurately reflect the direction of change in mucin expression in these analyses.

***Verification of the Proportional Hazards Assumption***

The key assumption of the Cox proportional hazards model is that the survival functions in the groups being evaluated remain proportional over time. Although this assumption can be validated visually (by ensuring Kaplan-Meier survival curves do not “cross”), the cox.zph function in R was also used to confirm proportionality for data that was analyzed using the Cox proportional hazards model. Using this method, p-values >0.05 indicate that the assumption of proportionality has been met.

E4. Khor CC, Do T, Jia H, Nakano M, George R, Abu-Amero K, Duvesh R, Chen LJ, Li Z, Nongpiur ME, Perera SA, Qiao C, Wong HT, Sakai H, Barbosa de Melo M, Lee MC, Chan AS, Azhany Y, Dao TL, Ikeda Y, Perez-Grossmann RA, Zarnowski T, Day AC, Jonas JB, Tam PO, Tran TA, Ayub H, Akhtar F, Micheal S, Chew PT, Aljasim LA, Dada T, Luu TT, Awadalla MS, Kitnarong N, Wanichwecharungruang B, Aung YY, Mohamed-Noor J, Vijayan S, Sarangapani S, Husain R, Jap A, Baskaran M, Goh D, Su DH, Wang H, Yong VK, Yip LW, Trinh TB, Makornwattana M, Nguyen TT, Leuenberger EU, Park KH, Wiyogo WA, Kumar RS, Tello C, Kurimoto Y, Thapa SS, Pathanapitoon K, Salmon JF, Sohn YH, Fea A, Ozaki M, Lai JS, Tantisevi V, Khaing CC, Mizoguchi T, Nakano S, Kim CY, Tang G, Fan S, Wu R, Meng H, Nguyen TT, Tran TD, Ueno M, Martinez JM, Ramli N, Aung YM, Reyes RD, Vernon SA, Fang SK, Xie Z, Chen XY, Foo JN, Sim KS, Wong TT, Quek DT, Venkatesh R, Kavitha S, Krishnadas SR, Soumittra N, Shantha B, Lim BA, Ogle J, de Vasconcellos JP, Costa VP, Abe RY, de Souza BB, Sng CC, Aquino MC, Kosior-Jarecka E, Fong GB, Tamanaja VC, Fujita R, Jiang Y, Waseem N, Low S, Pham HN, Al-Shahwan S, Craven ER, Khan MI, Dada R, Mohanty K, Faiq MA, Hewitt AW, Burdon KP, Gan EH, Prutthipongsit A, Patthanathamrongkasem T, Catacutan MA, Felarca IR, Liao CS, Rusmayani E, Istiantoro VW, Consolandi G, Pignata G, Lavia C, Rojanapongpun P, Mangkornkanokpong L, Chansangpetch S, Chan JC, Choy BN, Shum JW, Than HM, Oo KT, Han AT, Yong VH, Ng XY, Goh SR, Chong YF, Hibberd ML, Seielstad M, Png E, Dunstan SJ, Chau NV, Bei J, Zeng YX, Karkey A, Basnyat B, Pasutto F, Paoli D, Frezzotti P, Wang JJ, Mitchell P, Fingert JH, Allingham RR, Hauser MA, Lim ST, Chew SH, Ebstein RP, Sakuntabhai A, Park KH, Ahn J, Boland G, Snippe H, Stead R, Quino R, Zaw SN, Lukasik U, Shetty R, Zahari M, Bae HW, Oo NL, Kubota T, Manassakorn A, Ho WL, Dallorto L, Hwang YH, Kiire CA, Kuroda M, Djamal ZE, Peregrino JI, Ghosh A, Jeoung JW, Hoan TS, Srisamran N, Sandragasu T, Set SH, Doan VH, Bhattacharya SS, Ho CL, Tan DT, Sihota R, Loon SC, Mori K, Kinoshita S, Hollander AI, Qamar R, Wang YX, Teo YY, Tai ES, Hartleben-Matkin C, Lozano-Giral D, Saw SM, Cheng CY, Zenteno JC, Pang CP, Bui HT, Hee O, Craig JE, Edward DP, Yonahara M, Neto JM, Guevara-Fujita ML, Xu L, Ritch R, Liza-Sharmini AT, Wong TY, Al-Obeidan S, Do NH, Sundaresan P, Tham CC, Foster PJ, Vijaya L, Tashiro K, Vithana EN, Wang N, Aung T. Genome-wide association study identifies five new susceptibility loci for primary angle closure glaucoma. *Nat Genet* 2016; 48: 556-562.

E5. Marchini J, Howie B, Myers S, McVean G, Donnelly P. A new multipoint method for genome-wide association studies by imputation of genotypes. *Nat Genet* 2007; 39: 906-913.

E6. Marais S, Thwaites G, Schoeman JF, Torok ME, Misra UK, Prasad K, Donald PR, Wilkinson RJ, Marais BJ. Tuberculous meningitis: a uniform case definition for use in clinical research. *Lancet Infect Dis* 2010; 10: 803-812.

E7. van Laarhoven A, Dian S, Ruesen C, Hayati E, Damen M, Annisa J, Chaidir L, Ruslami R, Achmad TH, Netea MG, Alisjahbana B, Rizal Ganiem A, van Crevel R. Clinical Parameters, Routine Inflammatory Markers, and LTA4H Genotype as Predictors of Mortality Among 608 Patients With Tuberculous Meningitis in Indonesia. *J Infect Dis* 2017; 215: 1029-1039.

E8. van Laarhoven A, Dian S, Aguirre-Gamboa R, Avila-Pacheco J, Ricano-Ponce I, Ruesen C, Annisa J, Koeken V, Chaidir L, Li Y, Achmad TH, Joosten LAB, Notebaart RA, Ruslami R, Netea MG, Verbeek MM, Alisjahbana B, Kumar V, Clish CB, Ganiem AR, van Crevel R. Cerebral tryptophan metabolism and outcome of tuberculous meningitis: an observational cohort study. *Lancet Infect Dis* 2018; 18: 526-535.

E9. Shah TS, Liu JZ, Floyd JA, Morris JA, Wirth N, Barrett JC, Anderson CA. optiCall: a robust genotype-calling algorithm for rare, low-frequency and common variants. *Bioinformatics* 2012; 28: 1598-1603.

E10. McCarthy S, Das S, Kretzschmar W, Delaneau O, Wood AR, Teumer A, Kang HM, Fuchsberger C, Danecek P, Sharp K, Luo Y, Sidore C, Kwong A, Timpson N, Koskinen S, Vrieze S, Scott LJ, Zhang H, Mahajan A, Veldink J, Peters U, Pato C, van Duijn CM, Gillies CE, Gandin I, Mezzavilla M, Gilly A, Cocca M, Traglia M, Angius A, Barrett JC, Boomsma D, Branham K, Breen G, Brummett CM, Busonero F, Campbell H, Chan A, Chen S, Chew E, Collins FS, Corbin LJ, Smith GD, Dedoussis G, Dorr M, Farmaki AE, Ferrucci L, Forer L, Fraser RM, Gabriel S, Levy S, Groop L, Harrison T, Hattersley A, Holmen OL, Hveem K, Kretzler M, Lee JC, McGue M, Meitinger T, Melzer D, Min JL, Mohlke KL, Vincent JB, Nauck M, Nickerson D, Palotie A, Pato M, Pirastu N, McInnis M, Richards JB, Sala C, Salomaa V, Schlessinger D, Schoenherr S, Slagboom PE, Small K, Spector T, Stambolian D, Tuke M, Tuomilehto J, Van den Berg LH, Van Rheenen W, Volker U, Wijmenga C, Toniolo D, Zeggini E, Gasparini P, Sampson MG, Wilson JF, Frayling T, de Bakker PI, Swertz MA, McCarroll S, Kooperberg C, Dekker A, Altshuler D, Willer C, Iacono W, Ripatti S, Soranzo N, Walter K, Swaroop A, Cucca F, Anderson CA, Myers RM, Boehnke M, McCarthy MI, Durbin R, Haplotype Reference C. A reference panel of 64,976 haplotypes for genotype imputation. *Nat Genet* 2016; 48: 1279-1283.

E11. Howie B, Fuchsberger C, Stephens M, Marchini J, Abecasis GR. Fast and accurate genotype imputation in genome-wide association studies through pre-phasing. *Nat Genet* 2012; 44: 955-959.

E12. Beardsley J, Hoang NLT, Kibengo FM, Tung NLN, Binh TQ, Hung LQ, Chierakul W, Thwaites GE, Chau NVV, Nguyen TTT, Geskus RB, Day JN. Do Intracerebral Cytokine Responses Explain the Harmful Effects of Dexamethasone in Human Immunodeficiency Virus-associated Cryptococcal Meningitis? *Clin Infect Dis* 2019; 68: 1494-1501.

E13. Assarsson E, Lundberg M, Holmquist G, Bjorkesten J, Thorsen SB, Ekman D, Eriksson A, Rennel Dickens E, Ohlsson S, Edfeldt G, Andersson AC, Lindstedt P, Stenvang J, Gullberg M, Fredriksson S. Homogenous 96-plex PEA immunoassay exhibiting high sensitivity, specificity, and excellent scalability. *PLoS One* 2014; 9: e95192.
