## Supplementary Results for "*MUC5AC* genetic variation is associated with tuberculosis meningitis CSF cytokine responses and mortality"

***Validation of the proportional hazard assumption***

The proportional hazards assumption was met in the Vietnam discovery cohort (χ^2^= 0.80 with 1 degree of freedom [df]; p=0.37), the Vietnam validation cohort (χ^2^= 0.149 with 1 df; p=0.7), and the Indonesia validation cohort (χ^2^= 0.189 with 1 df; p=0.7)

***Results of Principal Component Analysis***

Principal component analysis of cytokine concentrations generated two factors, Factor 1 and Factor 2, representing highly correlated cytokines. None of the individual cytokines had uniqueness scores >70%, thus none were removed for individual analyses. Factor 1 had an eigenvalue of 5.46, explaining 74.1% of the variability in the data, with high loading values ( ≥0.56) for all cytokines. Factor 2 had an eigenvalue of 1.02, explaining 13.8% variability in the data. The cytokines TNF (0.59) and IFNγ (0.57) had the highest loading values for Factor 2. Some cytokines, including IL-6 (0.05) and IL-12 (0.01), had loading values near zero, whereas others (IL-2, -0.21; IL-4, -0.25; IL-5, -0.30; IL-10, -0.09; and IL-13, -0.32) had negative loading values for Factor 2, suggesting nearly all variability in Factor 2 was due to differences in concentrations of TNF and IFNγ.

***Supplementary Figure Legends***

**Figure E1: Comparison of additive, dominant and recessive models for the association between *MUC5AC* SNP rs28737416 and TBM mortality in the Vietnam discovery cohort.**

Kaplan-Meier curves displaying time to death in additive (**A**), dominant (**B**), and recessive (**C**) genotypic models. In the additive model, the homozygous recessive T/T genotype (solid, grey line) is compared to the heterozygous T/C genotype (dashed, gray line) and the homozygous dominant C/C genotype (dashed, black line). The dominant model compares a composite of the T/C and T/T genotypes (solid, grey line) to the C/C genotype (dashed, black line). The recessive model compares the T/T genotype (solid, grey line) to a composite of the T/C and C/C genotypes (dashed, black line). Displayed p-values represent comparison of the survival distributions by the log-rank test. The strongest association was seen for the dominant model, and thus a dominant model was used for all further analyses.

**Figure E2:** **Kaplan Meier curves comparing survival in a dominant allelic model of *MUC5AC* SNP rs28737416 in a subgroup of HIV-1 seronegative participants in the Indonesia validation cohort.**

Kaplan-Meier curves displaying time to death using a dominant allelic model comparing participants with either the T/T or T/C genotypes to those with the C/C genotype in HIV-1 seronegative participants in the Indonesia validation cohort are shown. Displayed p-values represent comparison of the survival distributions by the log-rank test.
