## Supplementary Table E1 for "*MUC5AC* genetic variation is associated with tuberculosis meningitis CSF cytokine responses and mortality"

**Table E1:** Evaluation of linkage disequilibrium between tagging SNPs in *MUC5B* and *MUC5AC* and previously identified functional SNPs in the *TOLLIP* gene.

|  | ***TOLLIP* SNPs** | |
| --- | --- | --- |
| ***MUC5AC* and *MUC5B* SNPs** | **rs5743854** | **rs3750920** |
| **rs28737416** | 0.02 | 0.01 |
| **rs2037089** | 0.00 | 0.01 |
| **rs7946008** | 0.02 | 0.04 |
| **rs6578810** | 0.81 | 0.28 |

R^2^ values representing linkage disequilibrium between index SNPs in *MUC5B* and *MUC5AC* and functional *TOLLIP* SNPs are shown.

Abbreviations: SNPs, single nucleotide polymorphisms
