## Supplementary Table E2 for "*MUC5AC* genetic variation is associated with tuberculosis meningitis CSF cytokine responses and mortality"

**Table E2:** **Evaluation of the association between *MUC5AC* SNP rs28738416 and 92 CNS cytokines among participants with TBM in the Indonesia validation cohort.**

| **Cytokine** | **Estimate** | **Std. Error** | **p-value** | **FDR** |
| --- | --- | --- | --- | --- |
| **IL-8** | -0.10 | 0.15 | 0.51 | 0.67 |
| **VEGFA** | -0.25 | 0.12 | 0.04 | 0.08 |
| **MCP3** | -0.73 | 0.31 | 0.02 | 0.06 |
| **GDNF** | -0.03 | 0.03 | 0.20 | 0.34 |
| **CDCP1** | -0.51 | 0.15 | 0.001 | 0.01 |
| **CD244** | -0.48 | 0.15 | 0.002 | 0.01 |
| **IL-7** | 0.03 | 0.07 | 0.68 | 0.77 |
| **OPG** | 0.04 | 0.13 | 0.75 | 0.81 |
| **LAP.TGFβ1** | -0.26 | 0.12 | 0.02 | 0.06 |
| **uPA** | -0.52 | 0.17 | 0.002 | 0.01 |
| **IL-6** | -0.28 | 0.32 | 0.39 | 0.55 |
| **IL-17C** | 0.01 | 0.05 | 0.86 | 0.89 |
| **MCP1** | -0.11 | 0.12 | 0.33 | 0.49 |
| **IL-17A** | -0.04 | 0.12 | 0.71 | 0.80 |
| **CXCL11** | -0.81 | 0.28 | 0.004 | 0.02 |
| **AXIN1** | -0.06 | 0.03 | 0.045 | 0.09 |
| **TRAIL** | -0.52 | 0.16 | 0.001 | 0.008 |
| **IL-20RA** | 0.01 | 0.008 | 0.46 | 0.62 |
| **CXCL9** | -0.18 | 0.17 | 0.31 | 0.47 |
| **CST5** | -0.02 | 0.02 | 0.34 | 0.50 |
| **IL-2RB** | 0.02 | 0.05 | 0.74 | 0.81 |
| **IL-1α** | -0.16 | 0.08 | 0.04 | 0.09 |
| **OSM** | -0.42 | 0.27 | 0.13 | 0.22 |
| **IL-2** | 0.05 | 0.06 | 0.43 | 0.60 |
| **CXCL1** | -0.10 | 0.18 | 0.56 | 0.72 |
| **TSLP** | 0.00 | 0.00 | 0.63 | 0.76 |
| **CCL4** | -0.53 | 0.20 | 0.009 | 0.03 |
| **CD6** | -0.60 | 0.16 | 0.0003 | 0.004 |
| **SCF** | -0.24 | 0.11 | 0.03 | 0.07 |
| **IL-18** | -0.66 | 0.20 | 0.0001 | 0.007 |
| **SLAMF1** | -0.51 | 0.16 | 0.002 | 0.01 |
| **TGFA** | 0.05 | 0.06 | 0.44 | 0.60 |
| **MCP4** | -0.23 | 0.11 | 0.04 | 0.09 |
| **CCL11** | -0.16 | 0.11 | 0.14 | 0.23 |
| **TNFSF14** | -0.29 | 0.17 | 0.10 | 0.17 |
| **FGF23** | 0.10 | 0.13 | 0.44 | 0.60 |
| **IL-10RA** | -0.14 | 0.04 | 0.002 | 0.01 |
| **FGF5** | 0.20 | 0.09 | 0.03 | 0.07 |
| **MMP1** | -0.41 | 0.23 | 0.09 | 0.15 |
| **LIFR** | 0.07 | 0.06 | 0.23 | 0.37 |
| **FGF21** | 0.08 | 0.13 | 0.55 | 0.71 |
| **CCL19** | -0.42 | 0.18 | 0.02 | 0.06 |
| **IL-15RA** | -0.24 | 0.06 | 0.0002 | 0.004 |
| **IL-10RB** | -0.35 | 0.12 | 0.004 | 0.02 |
| **IL-22.RA1** | 0.04 | 0.10 | 0.66 | 0.75 |
| **IL-18R1** | -0.33 | 0.16 | 0.04 | 0.08 |
| **PDL1** | -0.63 | 0.17 | 0.0003 | 0.004 |
| **βNGF** | -0.12 | 0.05 | 0.02 | 0.06 |
| **CXCL5** | -0.55 | 0.25 | 0.03 | 0.07 |
| **TRANCE** | -0.09 | 0.04 | 0.04 | 0.08 |
| **HGF** | -0.21 | 0.12 | 0.07 | 0.13 |
| **IL-12B** | -0.53 | 0.18 | 0.004 | 0.02 |
| **IL-24** | -0.03 | 0.05 | 0.63 | 0.75 |
| **IL-13** | 0.014 | 0.08 | 0.86 | 0.89 |
| **ARTN** | -0.004 | 0.01 | 0.75 | 0.81 |
| **MMP10** | -0.19 | 0.15 | 0.22 | 0.36 |
| **IL-10** | -0.48 | 0.21 | 0.02 | 0.06 |
| **TNF** | -0.44 | 0.18 | 0.02 | 0.05 |
| **CCL23** | -0.50 | 0.24 | 0.03 | 0.08 |
| **CD5** | -0.60 | 0.18 | 0.001 | 0.008 |
| **CCL3** | -0.51 | 0.21 | 0.01 | 0.05 |
| **Flt3L** | 0.04 | 0.08 | 0.65 | 0.75 |
| **CXCL6** | -0.24 | 0.22 | 0.29 | 0.44 |
| **CXCL10** | -0.07 | 0.10 | 0.48 | 0.64 |
| **EIF4EBP1** | -1.00 | 0.25 | 0.0002 | 0.004 |
| **IL-20** | 0.003 | 0.006 | 0.62 | 0.75 |
| **SIRT2** | -0.89 | 0.24 | 0.0003 | 0.004 |
| **CCL28** | -0.04 | 0.02 | 0.04 | 0.08 |
| **DNER** | 0.006 | 0.05 | 0.91 | 0.93 |
| **ENRAGE** | -0.57 | 0.24 | 0.02 | 0.05 |
| **CD40** | -0.61 | 0.18 | 0.001 | 0.007 |
| **IL-33** | 0.03 | 0.07 | 0.65 | 0.75 |
| **IFNγ** | -0.67 | 0.25 | 0.009 | 0.03 |
| **FGF19** | -0.12 | 0.10 | 0.23 | 0.37 |
| **IL-4** | -0.001 | 0.01 | 0.85 | 0.89 |
| **LIF** | -0.09 | 0.25 | 0.72 | 0.80 |
| **NRTN** | 0.00 | 0.00 | 0.60 | 0.75 |
| **MCP2** | -0.63 | 0.29 | 0.03 | 0.07 |
| **CASP8** | -0.76 | 0.21 | 0.0004 | 0.004 |
| **CCL25** | -0.33 | 0.12 | 0.007 | 0.03 |
| **CX3CL1** | -0.58 | 0.18 | 0.002 | 0.01 |
| **TNFRSF9** | -0.78 | 0.21 | 0.0002 | 0.004 |
| **NT3** | 0.02 | 0.03 | 0.65 | 0.75 |
| **TWEAK** | -0.08 | 0.08 | 0.28 | 0.43 |
| **CCL20** | -0.66 | 0.33 | 0.05 | 0.09 |
| **ST1A1** | -0.32 | 0.15 | 0.03 | 0.07 |
| **STAMPB** | -0.69 | 0.18 | 0.0001 | 0.004 |
| **IL-5** | 0.004 | 0.07 | 0.95 | 0.96 |
| **ADA** | -0.60 | 0.17 | 0.0004 | 0.004 |
| **TNFB** | -0.59 | 0.23 | 0.01 | 0.04 |
| **CSF1** | -0.14 | 0.08 | 0.07 | 0.13 |
| **BDNF** | -0.01 | 0.17 | 0.97 | 0.97 |
| **CD8A** | -0.63 | 0.27 | 0.02 | 0.06 |

Analysis of the association between the rs28738416 and CNS cytokine concentrations in validation cohort 2 using linear regression with ordinary least squares estimation and a Benjamini-Hochberg adjustment for multiple comparisons. Analyses were adjusted for age and Glasgow coma score.

Abbreviations: FDR, false discovery rate; STD, standard error; IL, interleukin
