## Supplementary figures and images for "*MUC5AC* genetic variation is associated with tuberculosis meningitis CSF cytokine responses and mortality"

### Supplementary Figure E1

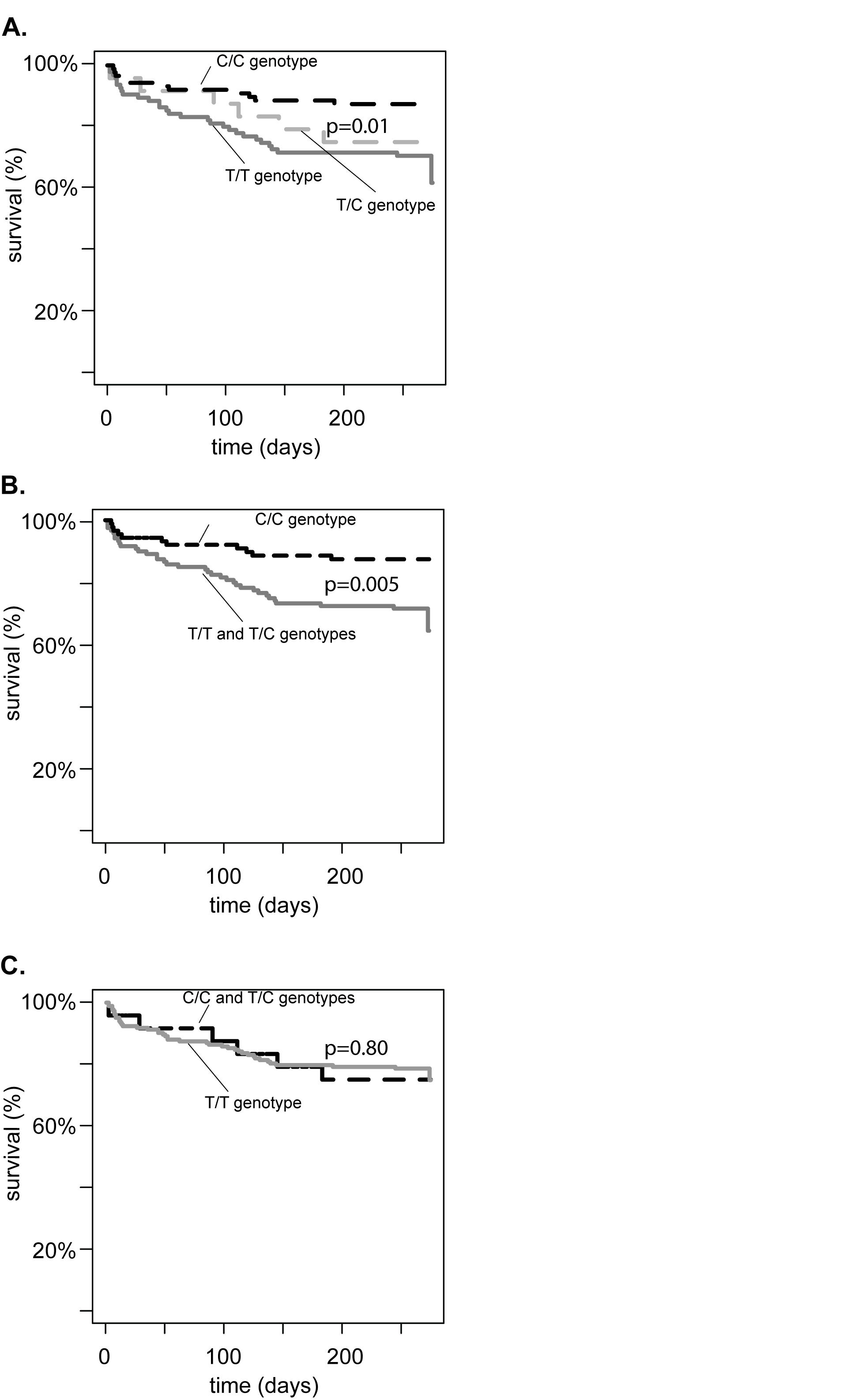

### Supplementary Figure E2

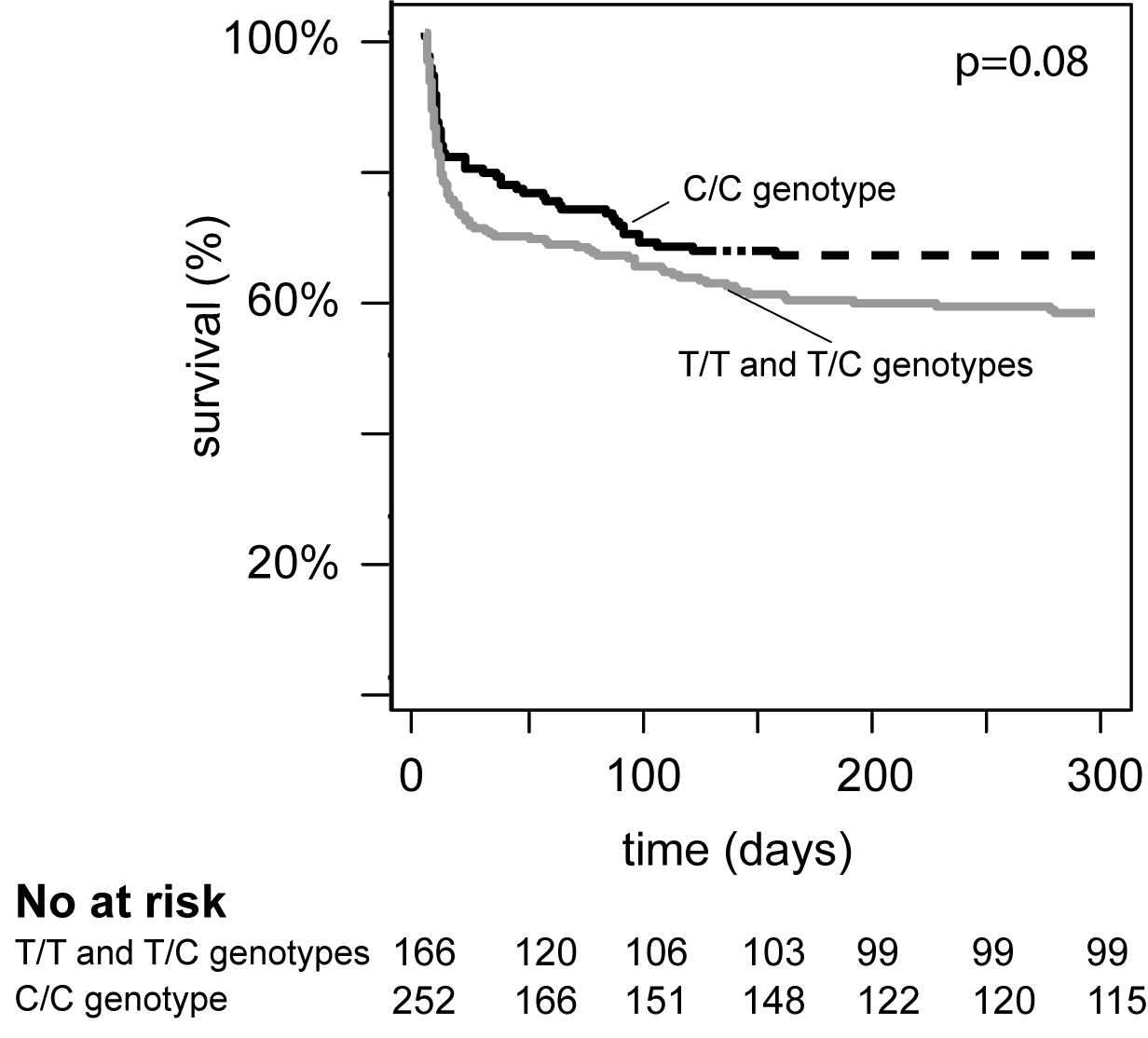
